# Developing a deep learning system to drive the work of the critical care outreach team

**DOI:** 10.1101/2020.07.07.20148064

**Authors:** Georgina Kennedy, John Rihari-Thomas, Mark Dras, Blanca Gallego

## Abstract

**Background:** Care of patients at risk of deterioration on acute medical and surgical wards requires timely identification, increased monitoring and robust escalation procedures. The critical care outreach role brings specialist-trained critical care nurses and physicians into acute wards to facilitate these processes. Performing this role is challenging, as the breadth of information synthesis required is both high and rapidly updating.

We propose a novel automated ‘watch-list’ to identify patients at high risk of deterioration, to help prioritise the work of the outreach team.

**Results:** This system takes data from the electronic medical record in real-time and creates a discrete tokenized trajectory, which is fed into a recurrent neural network model. These models achieve an AUROC of 0.928 for inpatient death and 0.778 for unplanned ICU admission (within 24 hours), which compares favourably with existing early warning scores and is comparable with proof of concept deep learning systems requiring significantly more input data.

**Conclusions:** Based on these results, we can conclude that it is technically feasible to build a set of predictive models that meet the needs of the critical care outreach role, based on a limited set of real-time clinical data.

## Background

### Clinical Setting

For a patient in an acute care setting, there are many complex and interrelated factors that affect their likely trajectory toward either recovery or deterioration. Prior to significant deterioration events, there are observable patterns in clinical features that indicate this change in acuity [1, 2, 3, 4]. These warning signs may be present as much as 48 hours prior to the adverse outcome [1], however they are often overlooked.

In addition, there is evidence that sub-optimal care (including delayed or missed interventions) in general hospital wards is a key contributing factor to both unplanned ICU admissions and preventable inpatient mortality [5, 6].

These factors have combined to drive the modern desire for tools and processes that can accurately highlight patients at risk of deterioration on the general wards such that interventions can be deployed sooner, improving both patient outcomes and resource utilisation. This commonly takes the form of an early warning score such as NEWS [7], which tracks physiological variables and raises an alert when they fall outside of acceptable limits.

It may also include the establishment of a critical care outreach team whose purpose is to integrate critical care skills of advanced assessment into the general care wards [8, 9]. This is a challenging role, requiring a rapidly updating awareness of events and patients across the whole hospital. In order to effectively prioritise their distributed workload, critical care outreach nurses and medical officers (CCON & CCOM) must synthesise information on a broader scale than is required of typical ward staff.

A physiological early warning score such as NEWS is intended to provide a trigger for emergency response, however the remit of the outreach role is broader than this — including the goal of identifying potential future deterioration in order to allow intervention prior to emergency onset. Risk models used to prioritise this work may therefore benefit from the inclusion of alternative risk factors such as pathology results or complex comorbidities. In addition to this, the reliance of existing models on vital signs indicators alone limits their capacity for automation in settings where these observations are not captured electronically.

### Technological Setting

There has been much interest in the development of deep learning models derived from electronic medical record (EMR) data. Deep-learning techniques are robust to heterogeneous, sparse and messy data, which are defining characteristics of the EMR. EMR data also fit naturally into recurrent neural network (RNN) architectures due to the discrete, episodic, time-series nature of the patient trajectory, which draws robust analogies to models of language. These language models have recently been expanded to account for the variable time intervals present in the patient record [10, 11, 12] by incorporating time-modulation gates or weightings for elapsed time.

Importantly, deep-learning techniques based on sequential tokens have the capacity to learn from rare events that would have insufficient predictive power in traditional models. Contextual embeddings such as the skipgram algorithm [13] transform high-dimensional one-hot encoded concepts into a lower-dimensional vector representation that can describe not only the exact event type, but also where the event type fits within a conceptual ‘neighbourhood’ [14]. This is done by learning a representation of events as they relate to adjacent events in the clinical trajectory — inferring that events that consistently appear in the same context will often contribute similarly to the patient’s overall risk profile.

Recurrent models have been developed from EMR data with high accuracy for diagnostic, phenotyping and prognostic purposes in diverse clinical domains. In particular, such systems have been demonstrated to perform well when used to predict inpatient mortality and ICU admission [10, 15, 16], which are the most important end-points for understanding short-term risk in a general patient population.

### Aim

The primary aim of this project is to investigate the feasibility of an automatically generated watch-list that provides outreach staff with an ordered list of patients most at risk of short-term deterioration. By analysing all available data in the medical record as it is generated, this list can supplement the clinical judgement of the CCON & CCOM and help them to proactively identify patients in need of early intervention to improve outcomes, avoid unnecessary or ineffective ICU admissions and reduce the risk of unexpected death.

The watch-list does not attempt to form a specific diagnosis nor prognosis but rather produces a priority list that can sit alongside clinical judgment. Users are therefore less tied to strictly explainable inference, requiring only a meaningfully calibrated relative risk. As such, we propose that it is a good candidate for piloting a real deep learning system in the clinical workflow. Preliminary user discussions suggest an openness to augment their workflow in this way, and a lower barrier for requiring exhaustive model scrutability due to the fact that the existing mental model for this role is so burdensome.

A significant limitation in this setting is the lack of any electronically recorded vital signs in the source data. All identified comparison deterioration models (both traditional [7, 17, 18, 19, 20, 21] and deep-learning [10, 15, 16]) rely on patient vital signs and physiological observations as key predictors. We are therefore also aiming to establish the viability of an alternative for predicting short-term patient deterioration where vital signs observations are not available. A study found that in settings where vital signs data are routinely documented using a mix of paper and electronic records, there are high levels of invalid and incomplete data [22], meaning that this limitation is sufficiently wide-spread that the automation of existing deterioration models would not be universally possible, and such an alternative is worth seeking.

## Methods

### Data

For this work, we used a dataset of hospital admissions from a metropolitan quaternary-care hospital in Sydney, Australia. The data were gathered retrospectively and approved for use by the target institution’s Human Research Ethics Committee.

All historical entries in the EMR were converted to discrete token values, based on their event type (admission/discharge, historical diagnosis, pathology results, medication administration, ward movement, surgical procedure or demography). These tokenized events were then concatenated to form a list of discrete values describing the patient’s historical trajectory that could be fed into the prediction model.

#### Example

Figure 1 shows an example of the inputs and prediction targets used to develop these predictive models. This example patient has two historical admissions (1a, 1b) prior to the current admission. Both historical admissions were for planned procedures, and include a mix of demographic and clinical tokens.

**Figure 1.**
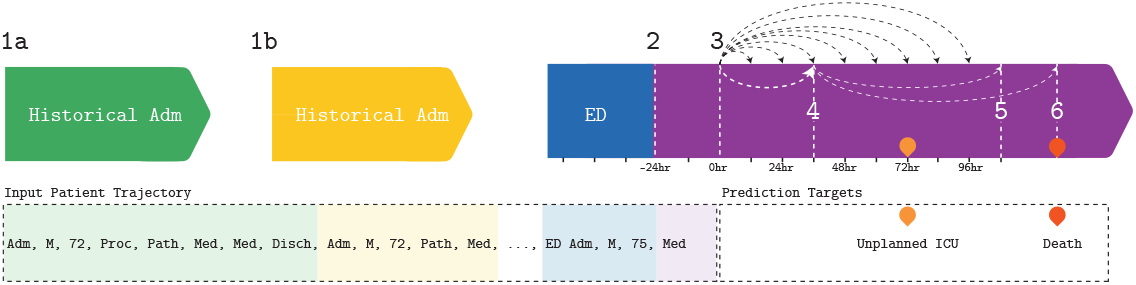
Example patient trajectory

In the target admission, the patient was admitted via the Emergency Department. Admission time (2) is the time that the patient was transferred to the medical wards. Prediction time (3) is set to 24 hours after admission time (t=0hr). All demographic and clinical tokens up to prediction time are included in the input data. Thus the input trajectory is an ordered list of all tokens occurring in any available historical admission(s), the patient’s ED stay, and the first 24 hours of the target admission.

### Targets

Events of interest are defined as in-hospital death and unplanned ICU admission, as a reduction in these events is the core premise supporting the establishment the critical care outreach team. There is no distinction made as to whether a death occurs in general wards, theatre or in the ICU.

No predictions are generated for patients in the ICU at the time of prediction, as they are already under the care of the core ICU team.

An ICU admission is classified as planned if it follows immediately from a surgical procedure, as there is no data available that specifically captures ICU admission intention. In the case that an admission to ICU direct from surgical theatres is indeed unplanned (i.e. due to unexpected in-theatre deterioration or adverse event), there is no intervention required from outreach staff, therefore the inability of the model to identify such cases is unlikely to be impactful.

Patients admitted directly to ICU are excluded from these models (363 admissions). In order to allow all states to be mutually exclusive and thus avoid the additional imbalance that would be introduced under a multiclass classifier accounting for death/ICU admission/both/neither, we train separate models for ICU admission and death risk.

Prediction time (*t* = 0*hr*) is set to 24 hours after a patient is admitted to general medical wards, either directly or via transfer from the emergency department. Prediction endpoints are measured at 12 hourly intervals, up to 4 days into the future (*t* = 0 + [12, 24, 36…96]*hours*).

### Data Preparation

In order to take advantage of the contextual embeddings that were initially developed for natural language processing (NLP) tasks, and as per prior deep learning work with EMR data [23], we converted each entry in the clinical database into token(s) of one of the following types: admission, discharge, pathology result, medication administration, ward movement, surgical procedure.

Pathology results and surgical procedure details contain continuous data types (numerical results, duration respectively), which cannot be handled by a straightforward contextual embedding model. These numerical values are therefore converted to decile results for each test or procedure type respectively. These tokens are then concatenated for each patient, with their associated time-delta since time of index admission, in order to describe their care trajectory, such as in Figure 1.

All data are inserted into the care trajectory at the time that they become available in the EMR. Ward movements, medication admission, pathology result, procedure and theatre movements are incorporated into the EMR in real-time. Some demography data are available at triage time, whilst some variables are input only at discharge. Coded diagnoses are not available in the EMR until some time after the time of discharge due to manual coding procedures. We therefore mask diagnosis codes associated with the target admission and only include historical diagnoses that end at least 3 days prior.

### Time-sensitive Concept Embedding

Before feeding such tokens into a deep-learning model, we must represent them numerically so that they may be used in the matrix algebra that forms the basis of the learning algorithm. An integer label for each distinct token in the vocabulary is insufficient for this purpose, as it implies an ordinality that does not exist and thus performs poorly. A one-hot encoding is possible, where each token is represented as an n-dimensional vector with a single ‘1’ corresponding to the specific term being described, and n is the number of distinct terms in the vocabulary. Such a representation typically leads to intractable calculations where n is a non-trivial number of available terms, and importantly does not take advantage of semantic similarity between terms (in this instance, perhaps a condition and its treatment are found to co-occur with sufficient frequency and particularity such that they may be treated similarly). These tokens were therefore transformed into a lower dimensional embedding space using a modification of the skipgram algorithm [13], which is a commonly used technique for assigning tokens a semantically-meaningful spatial representation.

Temporal and relational knowledge was encoded within the embedding by using a sampling function that was weighted inversely proportionally to both the timedelta between two events, and also whether or not the event occurred in the same admission. In equation 1, *w* is the weighted likelihood of selecting a particular pair of events as input to the skipgram algorithm, *s* is the distance between the two events by admission (for events in the same admission, *s=0*, for events in the admission immediately prior, *s=1* etc.), and *t* is the time interval between the two events in hours.

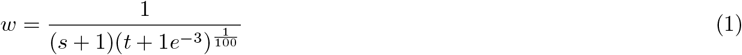

This weighting was then used to distribute the likelihood of sampling token pairs for inclusion in the embedding model. This is important because it allows the use of wide context windows in order to capture relationships between events occurring in rapid succession, as we want to preserve the strong relationship between temporally linked events (e.g. pathology results, where full test panels may return many results simultaneously) without introducing extraneous relationships between more loosely associated concepts captured within the same broad context window only incidentally due to the fact that there were no interposing events. This is a known challenge when learning low-dimensional embedding representations of clinical events [24, 25] without allowing for the time dimension. The effect of this decay factor is conceptually similar to the time-based dynamic windowing techniques in [26].

### Data Balancing

The targets of this model have a highly imbalanced distribution, which represents a significant challenge in the development of a useful model [27], with imbalances as skewed as 1 event in 160 for unplanned ICU admission and 1 in 180 for death within the shortest time-frames. We use a data augmentation strategy that allows the models to weight the loss functions appropriately and learn a more accurate representation of both the majority and minority classes.

Data augmentation is common in the domain of image processing tasks, where deep-learning has the longest history. It is typical to flip, rotate, skew, scale and mask portions of the input image in order to create multiple synthetic samples that retain the same class as the source, but allow a network to learn a more robust set of features that are less likely to over-learn idiosyncrasies related strictly to scale and positioning rather than the content of the image itself. Similarly, [28] applies window slicing and window warping strategies to provide synthetic samples from time-series data.

Following from these techniques, we implemented a data augmentation algorithm that can be applied to discrete time-series events such as those present in the EMR.

After copying trajectories and then randomly truncating the copies to 20-100% of their original length (by dropping the oldest events), time-series events were bucketed into 1 hour windows. 1 hour windows were chosen given the likelihood of meaningless time distinctions at any higher resolution based on an assumption of primarily manual data entry processes. Events within each of these 1-hour windows were then randomly shuffled and/or masked to create modulated patient trajectories which could be used to augment the input data. In the training set, each trajectory not including the target event was randomly augmented 4 times. Trajectories that included the target were augmented at a rate that was inversely proportional to the time to event (thus emphasising indicators of proximal deterioration), producing a balanced dataset. In the validation and test datasets, all trajectories were augmented 30 times, regardless of outcome.

### Final Models

The final model architecture was made up of three sub models that were trained jointly (Figure 2).

**Figure 2.**
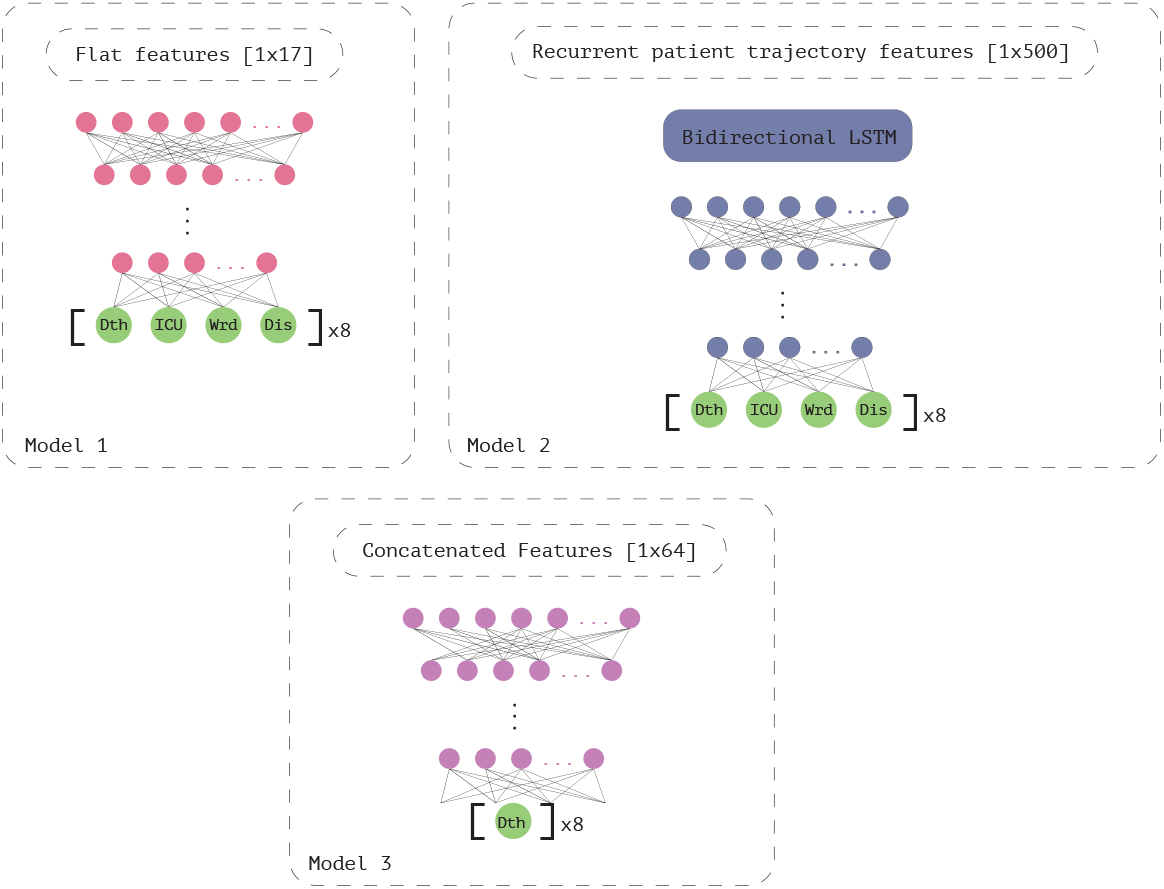
Model Architecture

*Model 1:* A flat set of features was created for each admission (see Table 1). These flat features were fed into a dense feed-forward network with a 4 dimensional output branch (Death, ICU, Discharge, Ward) for each of 8 time points (12, 24, 36, 48, 60, 72, 84 & 96 hours in the future). Terminal layer activation was set to Softmax, all prior layers had a LeakyReLU activation.

**Table 1.** Flat demography and historical summary features for each admission

| Feature | Range | Available (%) | Most common |
| --- | --- | --- | --- |
| Age (yrs) | 18-114 | 100 | 71 |
| Marital Status | 8 distinct | 96.2 | Married/partner |
| Aboriginal or Torres Strait Islander Ethnicity | 7 distinct | 98.7 | Neither |
| Insurance Status | 94 distinct | 100 | Medicare - overnight |
| Postcode of residence | 1497 distinct | 100 | Postcode of hospital |
| Country of birth | 220 distinct | 98.8 | Australia |
| Relative admission day | 0-2000 | 100 | 1703 |
| Admission hour | 1-23 | 100 | 7 (07:00hr) |
| Admission day of week | 0-7 | 100 | 3 (Tuesday) |
| Admission speciality | 47 | 98.6 | Emergency |
| Last discharge code | 9 distinct | 99.8 | 9 (discharge alive) |
| Days since last stay | 0-2000 | 62 | 2 |
| Last length of stay (LOS) | 0-475 | 62 | 0 |
| Historical total LOS | 0-592 | 62 | 0 |
| Historical average LOS | 0-237 | 62 | 0 |
| Historical ICU hours total | 0-2733 | 62 | 0 |
| Historical ICU hours mean | 0-1665 | 62 | 0 |

*Model 2:* The most recent 500 tokens in the patient trajectory were fed into a bi-directional LSTM layer, which then connected to a densely connected network, trained with the same 8 output branches as Model 1. Activations were also set as per Model 1.

*Model 3:* The 64 output variables from models 1 and 2 were concatenated into a single vector and used to train a densely connected network, with binary outcomes (i.e., death/∼death or ICU/∼ICU) at each of the target times.

#### Training Process

These models were trained jointly, meaning a single training batch was fed into models 1 and 2, with the resulting gradients back propagated, and then the output of this same batch was fed into model 3 and back propagated before moving onto the next training batch.

The models were trained on all 8 output times (12 to 96 hour forecasts), and then the loss function was modified to attend to the first 4 output times only and trained further in order to prioritise detection of imminent deterioration, whilst still allowing the model to learn from the more plentiful short to medium term deterioration end-points.

A 10% test set was held out with no processing applied until both ICU and Death model training was completed, with the remaining 90% used in a 5-fold cross validation process. At each fold, the training set was split into 80% training, 5% calibration and 15% validation sets.

### Calibration

A reference distribution of risks and uncertainty were produced by generating 300 predictions for each patient in the calibration set as per the validation data. We extend upon the binned calibration methods in [29] to transform the model output into a clinically-meaningful probability of deterioration.

For such short-term deterioration, it is a reasonable expectation that the proportion of patients deemed at low risk will far outweigh those at high risk. As such, instead of the fixed bin-widths in [29], we follow the argument in [30] for the use of unevenly spaced bins to generate measures of calibration quality to its logical conclusion and use these unevenly spaced bins to form the basis of the recalibration function itself.

This distribution was bucketed using a stick-breaking process at the quantiles 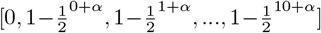 to generate scoring thresholds that appropriately reflected the far higher proportion of patients in low risk categories. A different *α* was selected for each category (correct, correct+72 hours, correct in admission) to reflect the different target distributions in the calibration set.

The risk score between 0 and 10 was then generated by comparing the predicted risk for each patient in the test set against these cutoff thresholds.

## Results

### Summary Statistics

Input data for these models included 192,883 hospitalisations, belonging to 92,802 adult patients (44.05% female), undergoing 117,658 surgical procedures over the period from June 2008 to June 2016. Patients had between one and 899 visits in the time period. Patients with 100 or more admissions (129 patients - all receiving regularly scheduled dialysis or rehabilitation treatments) were removed from the dataset so that they did not overwhelm the models, leaving a range of 1-99 admissions per patient (mean 2.08 *±* 3.92).

Patients had an average of 3864 *±* 7221 included clinical tokens at admission time. For admissions lasting more than 24 hours, 65 *±* 40 additional events were captured within the first day.

Admissions had one primary diagnosis and up to 44 associated comorbidities (mean 4.63 *±* 4.08). Every admission included by definition at least one ward movement (the ward to which the patient was initially admitted). Detailed summary statistics of the data can be found in the appendix.

### Endpoint Rates

Data imbalance is a well known challenge in the development of machine learning models. This is particularly relevant when the minority class is the class of interest, which is frequently the case in models that predict mortality, specific diagnoses or other important clinical end-points.

In the source admissions, there was an overall inpatient death rate of 1.53% and unplanned ICU admission rate of 3.22%. These rates change over the course of admission time, however, and drop drastically as the time windows become shorter (see Figure 3). At 24 hours after admission, the rate of death in the next 24 hours is 0.35% and for unplanned ICU admission it is 0.61%.

**Figure 3.**
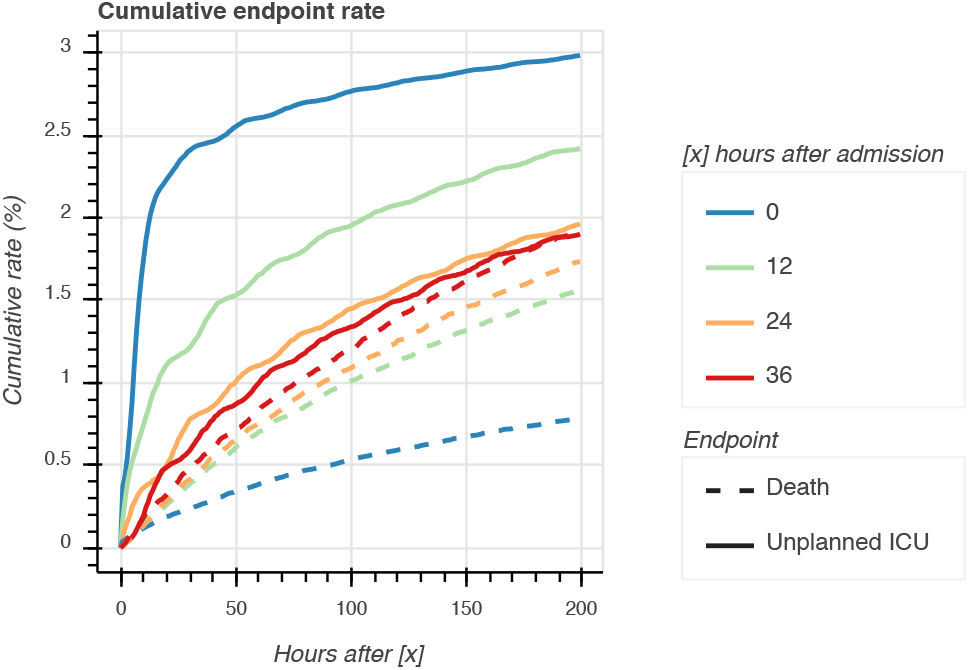
Endpoint rates in source data, relative to the number of patients still admitted at the given prediction point.

Unplanned ICU admission rates peak in the first day of admission and remain steady after that. Once an admission lasts more than 12 hours, the death rate becomes much higher. This is likely to represent the low death incidence within day-surgery admissions. From 12 hours onwards, the rate rises more gradually as the less severely ill patients are discharged. As death rates rise, unplanned ICU rates fall, which is indicative of an overall increase in acuity over time despite a decrease in instability.

### Reported Metrics

We report here metrics that test the output predictions against three measures:

1. A strictly correct forecast (model predicts endpoint within *t* hours, and this reflects accurately the presence of this endpoint within *t* hours).
2. A forecast that is correct with a clinically relevant tolerance. This tolerance is set to 72 hours (model predicts endpoint within *t* hours, and this reflects accurately the presence of this endpoint within *t* + 72 hours), to account for patients where similar response from outreach staff may be appropriate, given the desire for early intervention.
3. A forecast that is correct within the target admission (endpoint is predicted within *t* hours, and this is not necessarily accurate, however the endpoint of interest does occur prior to discharge). This gives a better sense of the true burden of false positives and false negatives on both patients and outreach staff.

In the example from Figure 1, there is an unplanned ICU admission at t=72hr, and the patient dies outside of the prediction window, but within this admission. At t=36hr (4), neither endpoint has occurred, so a prediction of false is strictly correct. Unplanned ICU admission does occur within 36+72 hours however (5), and therefore a prediction of ICU=true would be correct within the tolerance window and a prediction of death would be correct within the target admission.

For prediction use-cases with such high degrees of imbalance as those targeted by these models, with far more negative cases than cases of interest, reporting the area under the receiver operator curve (AUROC) alone can be highly misleading [31]. Despite this, it remains the most commonly reported statistic of model quality.

For this reason, we also report here the sensitivity and workup to detection ratio (WDR) for every prediction target. Model sensitivity is calculated as true positive predictions divided by all positive cases, or 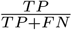. WDR is the inverse of the model positive predictive value, and provides the ratio of all positive predictions to all true positive predictions i.e 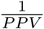, or 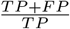.

Sensitivity is the key outcome measure from the perspective of at-risk patients. This is because a false negative corresponds to potential missed interventions and directly impacts their outcomes. WDR is the key metric for outreach staff however, as an increase in the burden of false positives will heavily reduce the usefulness of any predictive model, and may draw clinicians away from truly deteriorating patients. If balanced appropriately, these measures will result in the predictive model with the highest clinical utility.

Note that the AUROC frequently decreases as the tolerance increases, which is somewhat counter-intuitive, since a more permissive calculation could be expected to necessarily improve model performance. This is due to the fact that the tolerance does not only increase for the model predictions, but also for the model targets. Thus as the target event frequency increases the sensitivity calculation changes in both a positive and negative fashion, as more targets are correctly specified but more again are missed. This illustrates further the issue with reporting AUROC as the sole metric of model performance.

### Mortality Prediction

At 24 hours after admission, death within the following 24 hours was predicted with an AUROC of 0.928 (see Table 2 for all time points). This is higher than the baseline score NEWS [7] (0.89), however as outlined above, this measure alone is unlikely to tell the whole story of model utility. Note also that the NEWS baseline could not be replicated in the source data due to the unavailability of patient vital signs so is compared only to the AUROC as reported in the cited study.

**Table 2.** Area under the receiver operating curve for prediction within [x] hours, using data available 24 hours after admission time.

|  | [x] | 12 | 24 | 36 | 48 | 60 | 72 | 84 | 96 |
| --- | --- | --- | --- | --- | --- | --- | --- | --- | --- |
| **Death** |  |  |  |  |  |  |  |  |  |
|  | Correct forecast | 0.918 | 0.928 | 0.921 | 0.915 | 0.906 | 0.911 | 0.902 | 0.902 |
|  | Correct forecast with tolerance | 0.921 | 0.917 | 0.911 | 0.917 | 0.903 | 0.902 | 0.904 | 0.901 |
|  | Target within admission | 0.901 | 0.902 | 0.903 | 0.902 | 0.890 | 0.890 | 0.891 | 0.890 |
| **Unplanned ICU admission** |  |  |  |  |  |  |  |  |  |
|  | Correct forecast | 0.747 | 0.778 | 0.777 | 0.776 | 0.782 | 0.776 | 0.789 | 0.781 |
|  | Correct forecast with tolerance | 0.754 | 0.783 | 0.779 | 0.774 | 0.781 | 0.779 | 0.789 | 0.786 |
|  | Target within admission | 0.725 | 0.757 | 0.743 | 0.750 | 0.757 | 0.753 | 0.768 | 0.767 |

Figure 4 demonstrates the discriminative value of this model, i.e. that the output does indeed correspond to prediction of clinically meaningful risk. Although the sensitivity is poor at the earliest time point (due to the enormous class imbalance) later forecasts can be expected to correctly predict between a quarter and a third of patients who will deteriorate rapidly. Sensitivity drops as the tolerance increases to 72 hours, as there is now a higher proportion of target events. The workup to detection ratio decreases much more rapidly, however, demonstrating that the clinical burden of a false positive in this model is low, and that responding to a patient with even moderate risk is likely to be worthwhile.

**Figure 4.**
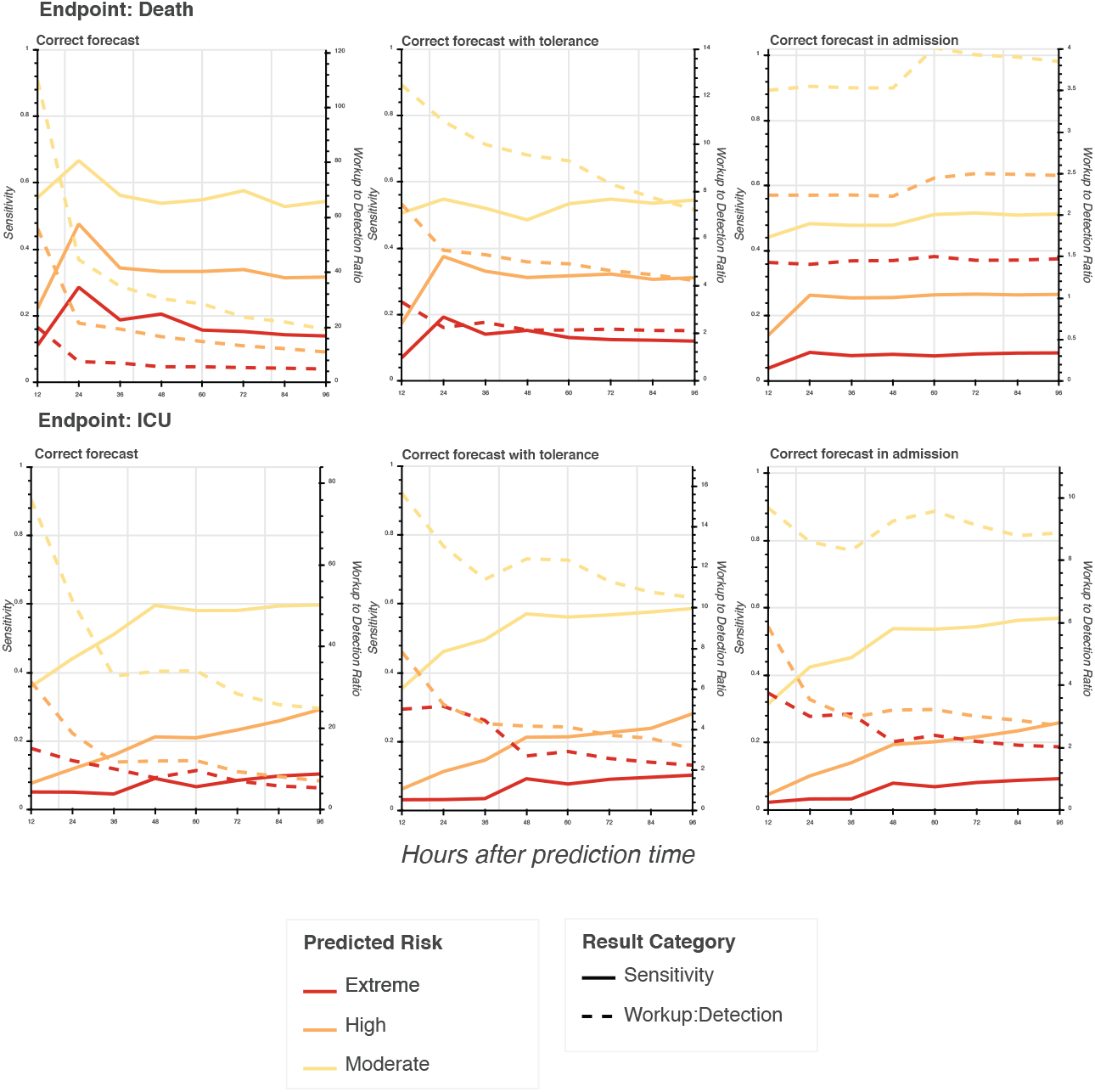
Mortality and unplanned ICU prediction — sensitivity and WDR of death prediction at future time points using data available at 24 hours after admission. For the purposes of risk stratification, extreme risk is here defined as a calibrated risk score of 6 or more, high risk as a score of 4 or 5, and moderate risk as a score of 2 or 3.

### Unplanned ICU admission

There is a significant difference between the AUROC of the mortality prediction models and the corresponding unplanned ICU admission models. This is likely to be due to the fact that ICU admission criteria are strongly coupled to vital sign triggers, and therefore a prediction model that does not include this data will underperform.

Despite this, from Figure 4, it remains possible to predict unplanned ICU admissions within the following 48 hours with a sensitivity of around 20% of all cases, and a corresponding WDR of 1 in 17. When allowing a 72 hour window of tolerance, a WDR of 1 in 12 gives up to 60% sensitivity, and therefore still represents a tool with meaningful clinical applications.

### Model Calibration

The raw results produced by this model had poor calibration, despite their good discriminative power, meaning that the probabilities output by the models could not be directly interpreted as the actual probability of the event occurring. This is typical of neural net techniques [30], which tend to be overconfident, or ‘sharp’ in their predictions.

There was a very low positive class count (not only proportionally, but also numerically) in the small calibration set. This meant that typical recalibration methods of isotonic regression [32] and Platt scaling [33] were ineffective (see Figure 5), and it also put techniques such as [34] out of reach.

**Figure 5.**
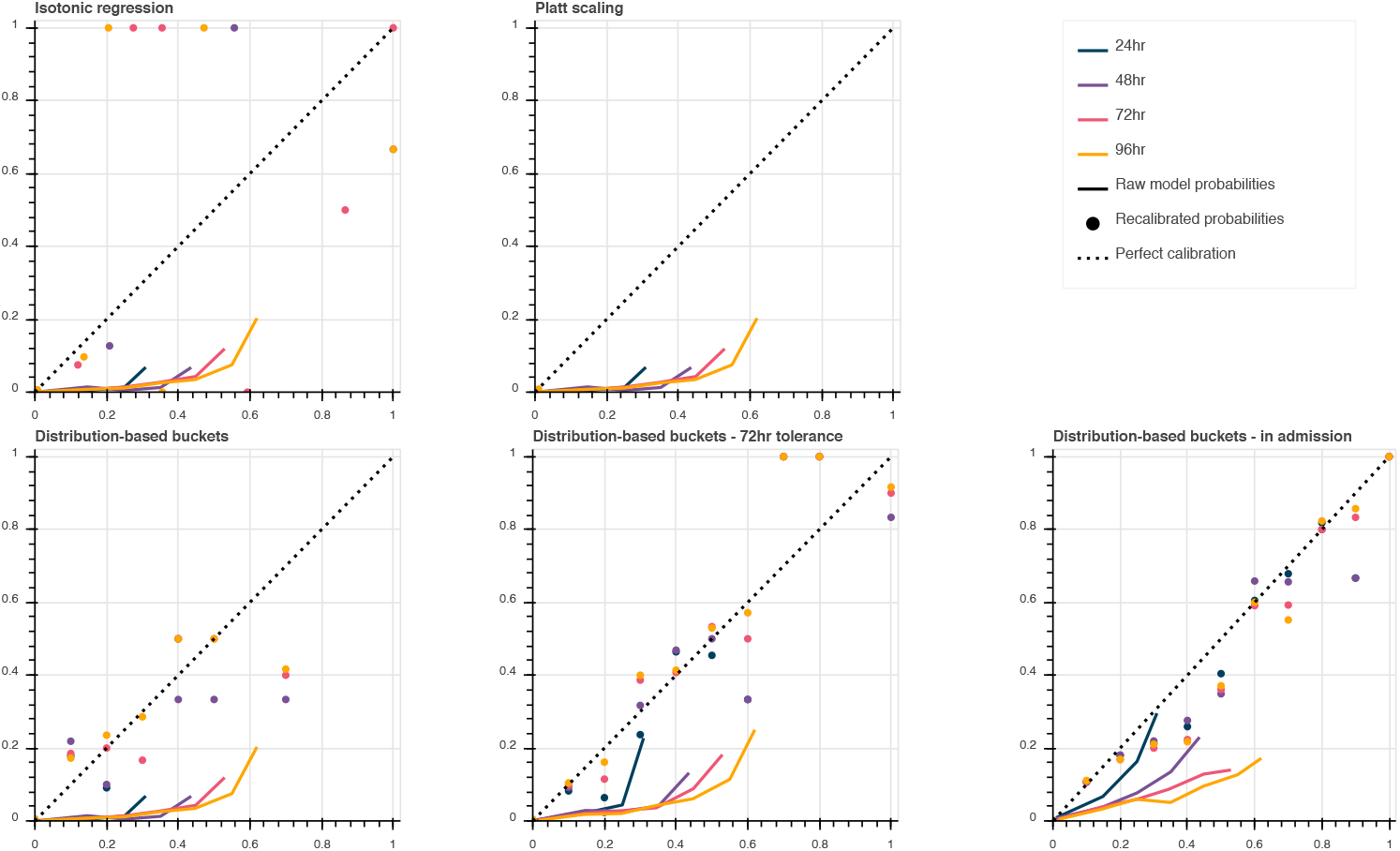
Recalibration techniques for death model predicted at 24 hours after admission. Note that Platt scaling reduces all probabilities to a single point close to the origin.

We find that the highest probability that we can assign to precise death forecasts is 40%, deaths within 72 hours of their forecast time have a maximal confidence of 80% and in-admission death has a maximum confidence of 90%. This matches the expectation that clinical trajectories are non-deterministic, particularly over the short term, but as the precise prediction time expands, confidence increases.

## Discussion

### Source Data Limitations

Scores or tools that target imminent patient deterioration typically aim to detect derangement of physiological signs and symptoms. This is based on the observation of predictable patterns of changes in patient vital signs prior to each of the relevant deterioration end-points cardiopulmonary arrest, unplanned ICU admissions and death [1, 2, 3, 35, 36].

Although a physiological early warning score (EWS) is used as a manual trigger of emergency response at the target institution [37], due to a lack of availability of vital sign data within the EMR, it is not currently possible to use such a score as the basis for a fully automated watch-list.

This, along with variable importance analyses in logistic regression models such as [19], serve to highlight the importance of vital sign data as the key element underpinning the vast majority of current best practice for prediction of inpatient deterioration. The limitation seen in our data is a realistic one, however, that should be considered for implementation of a fully automated system. It is characteristic of many EMR systems to serve the purposes of hospital administration first, and support clinically relevant data only where this aligns with the requisite billing and logistical goals, and/or where the clinical utility is high enough to justify the additional documentation burden above what can be provided with paper charts. Thus, it is unsurprising to observe in this data set that all theatre-based procedures are fully available in the clinical record, as they are not only billable, but also require the booking of resources from a central pool, compared with typical bedside procedures and nursing observations that go unrecorded for the inverse reasons.

This limitation in the breadth of input data is significant, however encourages a model that is built primarily around administrative data points, which are likely to be more reliably and consistently available in the EMR.

### Error Analysis

In order to understand the limitations of this model in these contexts, we ran the false positive samples with highest predicted risk (predicted death within 36 hours with a probability of 0.6 or higher but discharged alive) and the false negative samples with lowest predicted risk (died within 24 hours but death probability at 96 hours was lower than 0.2) through the LIME Text Explainer module [38]. LIME is an algorithm that provides insights into a ‘black-box’ model by learning a locally interpretable model that can explain which input data was most relevant to a given prediction.

In the word clouds in Figure 6, the size of a word corresponds to its weighted frequency as associated with each error type (false positives and false negatives). There is a clear pattern between the factors that contribute strongly to a prediction of high risk versus those contributing strongly to a low risk prediction. Lab results are generally indicative of a risk increase, where medications and medication-related tokens dominate lower risk predictions.

**Figure 6.**
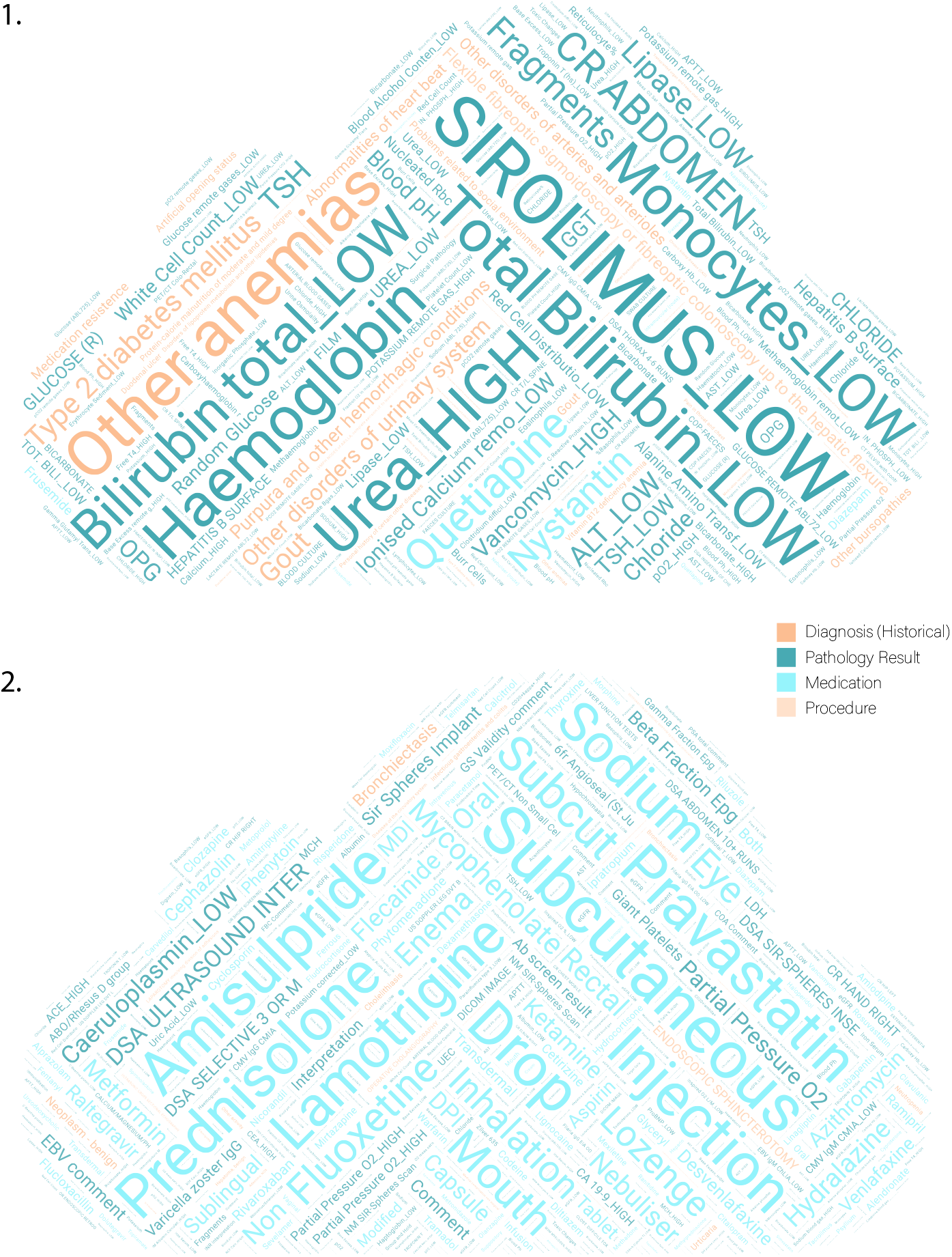
Word clouds demonstrating the most highly weighted terms for (1) false positive predictions and (2) false negative predictions.

For false negatives, most of these drug terms represent the highest-frequency tokens in the corpus. Their interpretation therefore is limited to the fact that they are evidence of a sort of regression to the mean, where these patients simply do not have enough distinctive data at the point of prediction to make an accurate risk assessment. Overall, despite having a comparable number of unique tokens, the medication terms each individually tend to have higher frequency than other token types. This holds true even when accounting for the repeated administration of medications, as these tokens on average each appear in more distinct patient trajectories than other event token types (excluding ward movement tokens).

In the list of terms contributing to false positives, there are numerous terms that may indicate that the patient has a complex history or is in a high-risk category, e.g. low white cell count, high blood urea, medication resistance, artificial opening status, sirolemus testing, low lipase. There are also, however, terms that either don’t have a sensible interpretation with respect to deterioration risk, e.g. low bilirubin, low blood alcohol content, Nystatin administration, or that are not sufficiently specific to make an informed interpretation of risk e.g. anaemia, sigmoidoscopy procedure, abdominal x-ray. This system is therefore insufficient to provide directed actions or interventions and its use must be limited to the prioritisation of attention.

### Congruence with Current Clinical Practice

The use of rapid response systems is intended to act as a safety net for deteriorating patients via the monitoring of a standardised subset of patient vital signs. It has, however, been argued that this drives nursing practice towards the detection of deterioration that is already well underway, as opposed to highlighting at-risk patients who are yet to go downhill [39]. By removing the reliance on vital signs, this model affords the capacity to move away from detection and into the realm of prediction.

Studies have also found that workloads and hospital work culture affect the likelihood of staff triggering rapid response calls according to the prescribed protocols [40]. Although calling criteria are nominally specified to allow triggering of the rapid response protocol based on clinical intuition alone (even when vital signs based criteria are not yet met) nursing staff who wish to act upon early signs of deterioration report themselves to be reluctant to do so in the face of potential criticism. This is true despite the fact that nursing intuition can preempt deterioration identified by vital signs alone [41]. A system that is able to provide contextualisation of such minor changes in patient state is therefore well placed to augment existing escalation protocols.

### Comparison Models

As a baseline, we present in Table 3 a selection of models that have been developed with the goal of detecting the early stages of short-term patient deterioration in a general ward population. Not all of these baselines can be compared directly to the models presented in this work due to the variability of endpoints and prediction times, giving instead an overview of the general targets and performances in existing models.

**Table 3.** Comparison to baseline models

| Model | Target Endpoint | Incl/Excl Criteria | Prediction Time | AUROC | Sens. | Spec. | Standardised WDR |
| --- | --- | --- | --- | --- | --- | --- | --- |
| *Traditional models* |  |  |  |  |  |  |  |
| NEWS [7] | In-hospital death (within 24hr) | Ex: Discharged before midnight of admission day; admitted directly to ICU | Time observations taken in medical | 0.89 | - | - | - |
|  | Unplanned ICU (within 24hr) |  |  | 0.86 | - | - | - |
|  | Cardiac arrest (within 24hr) |  |  | 0.72 | - | - | - |
|  | Combined 24hr deterioration |  |  | 0.87 | - | - | 31.5 |
| Alvarez et al [17] | Resuscitation events and death | Inc: Adult patients admitted to internal medicine ward or ICU. Ex: admitted directly to surgery; DNR order at admission; obstetrics admission; events on first day of admission | Daily prediction | 0.85 | 0.52 | 0.94 | 35.6 |
| Churpek et al (a) [18] | Cardiac arrest (in admission) | Inc: Adult patients with documented vital signs | Every 8 hours | 0.88 | - | - | - |
|  | Unplanned ICU (in admission) |  |  | 0.77 | 0.54 | 0.90 | 55.6 |
|  | Cardiac arrest (within 24hr) |  |  | 0.88 | 0.65 | 0.93 | 33.2 |
|  | Unplanned ICU (within 24hr) |  |  | 0.76 | - | - | - |
| Churpek et al (b) [19] | Combined 8hr deterioration | Inc: Adult patients with documented vital signs | Every 8 hours | 0.80 | 0.50 | 0.93 | 42.8 |
| Kipnis et al [20] | Combined 12hr deterioration | Inc: Adult patients. Ex: out of network transfers; childbirth admissions, 'comfort care only' orders. | Hourly | 0.82 | 0.49 | 0.92 | 49.5 |
| Green et al [21] | Combined 24hr deterioration | Inc: All admissions. | At time of vital sign observation | 0.80 | 0.50 | 0.90 | 59.9 |
| *Deep learning models* |  |  |  |  |  |  |  |
| Rajkomar et al [10] | In-admission death | Inc: Length of stay > 24hr; adult patients | 24hr after admission | 0.95 | - | - | - |
| CCO watch-list (this work) | Unplanned ICU (within 48hr) | Inc: Length of stay > 24hr; adult patients, fewer than 100 visits, not admitted directly to ICU | 24hr after admission | 0.77 | 0.50 | 0.88 | 71.2 |
|  | In-hospital death (within 24hr) |  |  | 0.93 | 0.47 | 0.97 | 21.3 |

**Notes:** Where more than one result available for same end-point, result with highest AUROC is reported. Where more than one prediction time is available, most clinically relevant prediction time for that end-point is reported. Where multiple cutoff points are available, sensitivity and specificity are reported as per review paper [42]. Workup to detection ratio is only reported where it is possible to standardise this measure to a fixed reference prevalence rate. Reference rate has been set to 0.35% for all WDR calculations, setting sensitivity in range ~50% per [42]. For NEWS, fixed sensitivity/specificity in target range not available. WDR instead calculated from EWS efficiency curve.

Note that it is only possible to compare WDR to baselines reported in different populations if a fixed incidence rate is chosen in order to standardise this measure. Where it was possible to make this calculation, the fixed rate was set to 0.35%, which is the death rate within 24 hours in this population, per section.

The traditional models were identified from a recent review paper that is closely aligned with the target use-case [42] in addition to the NEWS model [7], which is a highly cited and widely implemented early warning score that forms the basis for comparison for many similar works.

In order to capture potential deep learning baselines, the reference list of two systematic reviews [43, 44] were filtered to identify EMR-based patient deterioration prediction models. General deterioration endpoints not applicable to the CCON/CCOM role were excluded, e.g. readmission, death other than short-term, or studies only applicable to patients already within the ICU. Notably, many deep learning models do not fit our use-case as they either predict only inpatient or longer-term mortality e.g. [45, 46, 10], target a specific morbidity such as congestive heart failure or sepsis e.g. [47, 48] or are developed using data for patients already admitted to the ICU e.g. [49, 50] (largely due to the wide utilisation of the freely-available MIMIC-III database [51]). [10] was retained as the deep learning baseline, as it is closest to meeting the target use-case. Interestingly, this reference uses the NEWS model as a mortality baseline, despite the fact that NEWS was developed to detect 24-hour mortality where the deep learning model predicts inpatient mortality.

This summary of baselines exposes a number of issues with the comparison of such predictive systems. In particular, the precise definition of endpoints is inconsistent. We also note that all mortality endpoints reported here are for in-hospital mortality only, i.e. they are unable to report full mortality as an endpoint due to the lack of data linkage and potential loss to followup. Only Kipnis et al [20] have access to network-level data linkage, but this is not utilised as a primary endpoint. Rajkomar et al [10] go further by redefining readmission to include only readmission to the same institution. The availability of linked data as per [52] would provide additional insight and allow expansion of these models to include identification of patients at the end of life.

### Data Processing

Many clinical prediction scores rely on highly regulated data collection that may not reflect existing clinical processes, thus requiring additional data entry or hand calculation. Our noisy dataset reflects true practice and availability, with pre-processing limited to routines that can be performed with no human input. Within this preprocessing of data, we do not attempt to normalise the labelling of medications and pathology — e.g. different spellings are present for the same test across different panels — instead, allowing contextual embeddings to handle this noise. We do this on the assumption that the more hands off we are in data preparation, the more robust the results will be to changing practice and the lower effort required by the end-users. We also do not make any effort to handle multiple recordings at the same time, or detect outliers.

Because we rely on the naturalistic data ecosystem, rather than one requiring abstraction, we assume that we are reducing errors caused by hand calculations or operational error, and robust to errors preexisting within the EMR. The trade-off with this strategy is that we cannot expect these models to achieve generalisation in a new setting without re-training to accommodate local vocabularies and idiosyncrasies of data entry. An external validation study will therefore require translation of the entire model pipeline, rather than transfer and mapping of only the model inputs themselves.

### Calibration Measure

It is not feasible to calculate the Hosmer-Lemeshow statistic of calibration for this model due to the large sample size and excessive degrees of freedom [53, 54]. Alternative calibration statistics were reviewed for their applicability such as [55], however were found to be unsuitable due to their focus on density. This makes sense for many use-cases, where it is valuable to prioritise areas of the calibration curve that represent the majority of samples, however in this situation it is not suitable, as the differences between probabilities at the low end of the risk scale are not clinically meaningful. Instead, the differences in the most sparse regions must be prioritised — outreach staff may be expected to treat patients at 80% risk quite differently to those at 90% risk, despite there being very few patients in those risk categories, where their response will differ very little for patients at 10% risk vs. 20% risk.

This knowledge-based interpretation of the utility of a model’s calibration cannot be quantified without some parameters set by target users a priori.

## Conclusion

Based on these results, we can conclude that it is technically feasible to build a set of predictive models that meet the needs of the critical care outreach role, based on a limited set of real-time clinical data. These models compare favourably with the current practice of using physiological early warning scores to highlight deteriorating patients when compared numerically in terms of accuracy, AUROC and workup to detection ratio, although there remains a significant amount of work to successfully implement them in practice.

## Data Availability

The data input for the current study are not publicly available. Due to reasonable privacy and security concerns, the data cannot be distributed to researchers other than those granted access via the Human Research Ethics Committee of the target institution.

### Appendix

#### Summary Statistics

The dataset used as input for this study comprised 192,883 admissions, shared by some 92,802 patients. See Table 4 and Figure 7 for detailed summary statistics. It is important to notice the extreme skew in the distribution of some data elements across patient records, in particular pathology and medication records, where the mean number of records per patient outstrips the mode by several orders of magnitude. This lack of uniformity in the richness of input data is a key challenge for any predictive task.

**Table 4.** Population Statistics

| Item | Count | Mode | Mean | St.Dev. |
| --- | --- | --- | --- | --- |
| Patients | 92802 |  |  |  |
| Male | 51844<br>(55.9%) |  |  |  |
| Female | 40958<br>(44.1%) |  |  |  |
| Age |  | 71 | 59.96 | 20.34 |
| Admissions | 192883 |  |  |  |
| Admissions per patient |  | 1 | 2.07 | 3.92 |
| All diagnoses | 892629 |  |  |  |
| Primary diagnoses | 192863 |  |  |  |
| Comorbid diagnoses | 699766 |  |  |  |
| Diagnoses per patient |  | 1 | 9.62 | 17.73 |
| Diagnoses per admission |  | 1 | 4.63 | 4.08 |
| Distinct diagnoses per patient |  | 1 | 7.12 | 8.61 |
| Medication administration events | 12524922 |  |  |  |
| Medication events per patient |  | 0 | 134.96 | 506.50 |
| Per patient with $\geq 1$ event | | 1 | 252.25 | 670.74 |
| Pathology results | 41871520 |  |  |  |
| Pathology results per patient |  | 0 | 451.19 | 1221.09 |
| Per patient with $\geq 1$ result | | 2 | 479.85 | 1253.80 |
| Surgical procedures | 117658 |  |  |  |
| Surgical procedures per admission |  | 0 | 0.61 | 1.36 |
| Surgical procedures per patient |  | 0 | 1.27 | 2.68 |
| Per admission with $\geq 1$ procedure | | 1 | 1.92 | 1.82 |
| Per patient with $\geq 1$ procedure | | 1 | 2.85 | 3.42 |
| Ward movements | 676193 |  |  |  |
| Ward movements per admission |  | 1 | 3.51 | 3.26 |
| Ward movements per patient |  | 2 | 7.29 | 11.12 |
| Time between admissions (days) |  | 2 | 105 | 238 |
| Length of stay (hours) |  | 3 | 98 | 239 |
| Length of stay for stays $\geq 24$h | | 27 | 196 | 316 |

**Figure 7.**
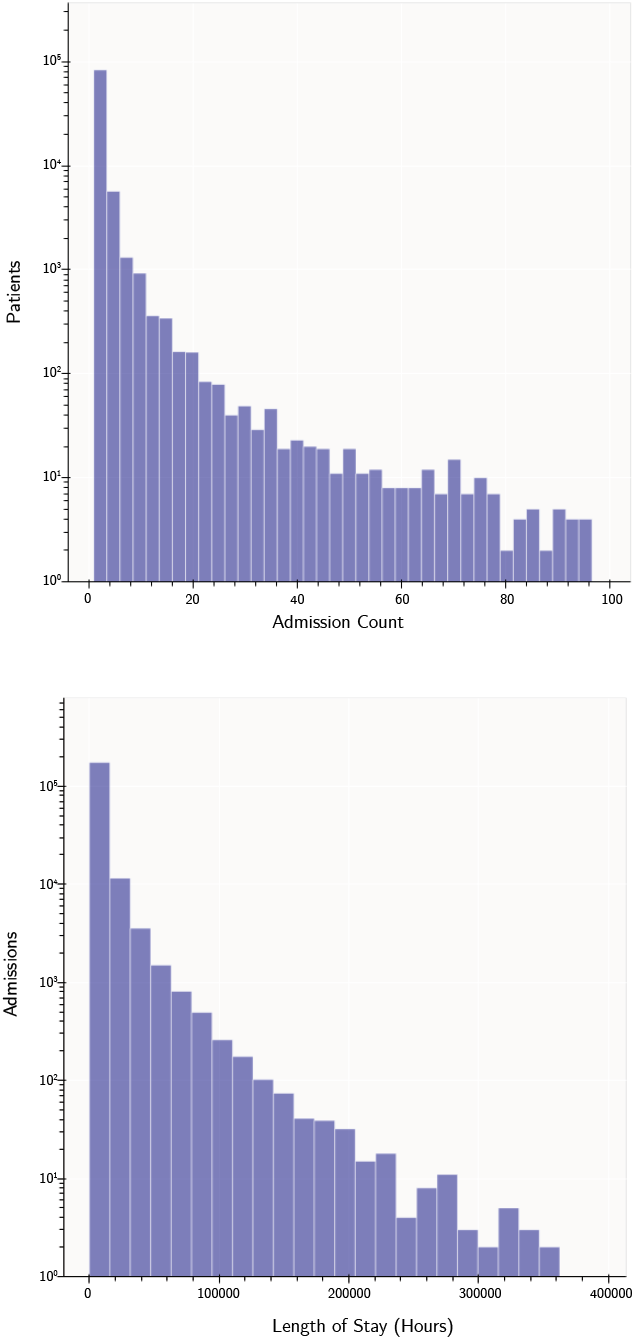
Distributions: admissions per patient, length of stay

## Funding

This research is supported by an Australian Government Research Training Program (RTP) Scholarship.

### Abbreviations

AI: Arti_cial intelligence
AUROC: Area Under the Receiver Operating Characteristic curve
CCO: Critical Care Outreach
CCOM: Critical Care Outreach Medical O_cer
CCON: Critical Care Outreach Nurse
EMR: Electronic Medical Record
EWS: Early Warning Score
ICU: Intensive Care Unit
LIME: Local Interpretable Model-Agnostic Explanations
LSTM: Long Short Term Memory unit
MET: Medical Emergency Team
NEWS: National Early Warning Score
NLP: Natural Language Processing
RNN: Recurrent Neural Network
WDR: Workup to Detection Ratio

## Availability of data and materials

The source data for this work cannot be made publicly available due to restrictions of the ethics approval. Code that applies much of this processing pipeline to the publicly available MIMIC III dataset [51] can be found at https://github.com/CBDRH/PaTMan

## Ethics approval and consent to participate

This study was approved as ethics application HREC/15/SVH/403 under the St Vincent’s Hospital Human Research Ethics Committee. This approval was granted on the 10^th^ of August, 2016 and is valid until the 2^nd^ of August, 2021.

## Consent for publication

Not applicable.

## Authors’ contributions

GK: Data analysis, model development, model architecture conception, manuscript preparation; JRT: Clinical guidance, manuscript input and review; MD: Technical guidance, manuscript input and review; BG: Overall guidance and direction of model development, manuscript input, project supervision

## Acknowledgements

Not applicable.

## Authors’ information

Not applicable.

